# Assessing the impact of frailty on statin prescriptions among older stroke survivors with and without diabetes in Malaysia

**DOI:** 10.1101/2025.05.05.25326965

**Authors:** Wei Jin Wong, Mei Shin Yau, Kit Mun Tan, Tu Nguyen

## Abstract

**Background:** Statins are crucial in secondary prevention for patients post strokes. Frailty may influence prescribing decisions for statins, and the interaction between frailty and diabetes in the context of statin use remains unexplored.

**Aim:** This study aimed to examine the relationship of frailty and statin prescriptions in older participants with stroke, and to compare statin prescription rates and the impact of frailty on statin prescriptions between participants with and without type 2 diabetes.

**Methods:** A cross-sectional study was conducted in older patients with ischemic stroke in a tertiary hospital in Malaysia. Frailty assessments prior to discharge from hospital were determined using the Clinical Frailty Scale version 2.0. Odds ratios (ORs) were estimated from logistic regression models to examine the relationship between frailty and the prescription of statins.

**Results:** There were 282 participants (mean age 80.8, SD 6.3), 132 with diabetes and 162 were female. The mean CFS score was 6.1 (SD 1.1) in all participants, 6.0 (SD 1.1) in participants without diabetes, and 6.2 (SD 1.0) in participants with diabetes (p=0.099). The percentages of frailty (defined with a CFS ≥4) were 97.2% in all participants, 96.0% in those without diabetes vs 98.5% in those with diabetes (p=0.290). Statin was the most commonly prescribed medication at discharge (69.0%), followed by antiplatelets (65.5%), calcium channel blockers (38.4%). There was no significant difference on statin prescription rates between participants with and without diabetes (71.0% vs. 67.3%, p=0.508). Increased CFS score was significantly associated with reduced odds of receiving statins in all participants (adjusted OR 0.64, 95% CI 0.46-0.88), and in participants without diabetes (adjusted OR 0.54, 95% CI 0.33–0.89), but not in participants with diabetes (adjusted OR 0.73, 95%CI 0.46–1.17).

**Conclusion:** Frailty was associated with reduced odds of receiving statins in the study population. The differences in the relationship between frailty and statin prescriptions among participants with and without diabetes may suggest a personalized approach in secondary prevention for older patients after strokes. Future studies are needed to understand prescribers’ perspectives, aiding in the development of personalized healthcare for older individuals.

## Introduction

Stroke continues to be a leading source of mortality and morbidity worldwide, with the bulk of the global burden in lower-income and lower-middle-income countries.^1^ There are two main types of stroke, ischemic stroke and hemorrhagic stroke. With age being an irreversible risk factor for stroke, older persons are more vulnerable to stroke and its long-term effects. For older stroke survivors, managing cardiovascular risk factors such as hyperlipidaemia is a crucial strategy in reducing the likelihood of subsequent cardiovascular events and further complications. Strategies can include non-pharmacological steps like lifestyle interventions and pharmacological options such as the use of lipid lowering therapies like statins. Statins, also known as hydroxymethylglutaryl-coenzyme A (HMG-CoA) reductase inhibitors, are primarily used for their lipid-lowering effects and cardiovascular benefits. Statins are recommended by the American Heart Association and American Stroke Association^2^ for the prevention of recurrent stroke in patients post ischemic strokes.

Frailty is a condition characterized by decreased physiological reserve and increased vulnerability to adverse health effects.^3^ Frailty has also been linked to a higher incidence of future cardiovascular events.^4^ A study of 15753 participants with type 2 diabetes from the UK Biobank showed that both pre-frailty and frailty were associated with a higher risk of cardiovascular events, including strokes.^5^ Therefore, the presence of frailty in patients with diabetes could significantly increase the risk of recurrent strokes. As such, the impact of frailty on cardiovascular diseases including stroke is an important research area.^6^ However, for older stroke survivors with frailty, this continues to be an under-explored area. With reports of increasing prevalence of frailty among older people, its importance and influence are growing, particularly in how it impacts care. Frailty may influence clinical decisions, including the prescriptions of medicines like statins.

Malaysia is a country in the Southeast Asia that is undergoing an epidemiological transition.^7^ The increasing number of older population, and with it frailty, presents many challenges for the healthcare system. Furthermore, the prevalence of type 2 diabetes in older adults in Malaysia is high.^8^ People with diabetes have an increased risk of stroke and post-stroke complications. For stroke care, the combination of a growing number of older people, together with the reported high prevalence of poorly controlled cardiovascular risk factors like blood pressure control in people with diabetes^9^ further highlights the magnitude of the challenge at hand. There is a need to better understand how these influence strategies like statin prescription as part of stroke care for the older person. Statins have been reported to be underutilized for secondary prevention and there is limited data on the relationship of frailty and the use of lipid lowering therapies in this population.^4^ The interaction between frailty and diabetes in the context of statin use also remains unexplored. To help improve stroke care for older persons, it underscores the need for further incorporation of geriatric-related issues like frailty and how it relates to other risk factors like diabetes.

Therefore, this study aimed to examine the association of frailty and the prescriptions of statins in older participants with stroke, and compare statin prescription rates and the impact of frailty on the prescriptions of statins between participants with and without type 2 diabetes

By examining these factors, this study seeks to enhance understanding of how frailty and diabetes influence clinical decision making such as prescribing habits.

## Methods

We used data from an observational study on the prevalence of pre-stroke frailty and its impact on outcomes in 384 older patients (aged 65 years or over) in Malaysia from 2016 to 2020. This database also had data on frailty status of the patient post-stroke and prior to discharge from hospital. Further details of the study was described elsewhere.^10^ Ethics approval was obtained from the Medical Research Ethics Committee of the University Hospital (MEC No: 201312 – 0636). For the purposes of this analysis, only patients with ischaemic stroke (n=336) were included.

Participants’ characteristics such as the socio-demographics, comorbidities including diabetes, and post-stroke function, were obtained from electronic medical records. All cases of diabetes in this study were type 2 diabetes. Disability at discharge was defined with a Modified Rankin Scale (MRS) of 3 or above.

Frailty was defined according to the Clinical Frailty Scale (CFS) version 2.0. The CFS v2.0 score ranges from 1-9 and a cut-point of 4 was applied to define frailty.^11^

Information on prescribed medications was obtained from the hospital electronic discharge summary system by a trained research assistant. Information collected included number of medicines, name of medicine together with dose, frequency and route. Statin use was defined as the prescriptions of any statin medication. In this study, the types of statins that were prescribed included atorvastatin, rosuvastatin, and simvastatin. Polypharmacy was defined as the presence, on prescriptions at hospital discharge, of five or more medications.^12^

### Statistical analysis

Continuous variables are presented as mean and standard deviation (SD), and categorical variables as frequency and percentage. Comparisons between participants with and without diabetes were assessed using the Chi-square test or Fisher’s exact test for categorical variables and Student’s t-test or Mann-Whitney test for continuous variables. Two-tailed p-values < 0.05 were considered statistically significant.

To examine the relationship between frailty and the prescription of statins, odds ratios (ORs) were estimated from logistic regression models, unadjusted and adjusted for pre-defined covariates that may affect statin prescriptions, including age, sex, comorbidity burden (the Charlson Comorbidity Index), dyslipidemia, history of previous stroke, polypharmacy, and disability at discharge (MRS ≥3). Frailty was treated as a continuous variable (CFS score). Results are presented as ORs and 95% confidence intervals (CIs) for all participants, and for those with and without diabetes.

Analysis of the data was performed using SPSS 29.0.

## Results

A total of 282 participants with data of frailty assessment and medication prescriptions upon discharge were included (150 without diabetes, 132 with diabetes). They had a mean age of 80.8 (SD 6.3) years and 57.4% were female. Table 1 presents the participant general characteristics, stratified by diabetes. The mean CFS score was 6.1 (SD 1.1) in all participants, 6.0 (SD 1.1) in participants without diabetes, and 6.2 (SD 1.0) in participants with diabetes (p=0.099). The percentages of frailty (defined with a CFS ≥4) were 97.2% in all participants, 96.0% in those without diabetes vs 98.5% in those with diabetes (p=0.290).

**Table 1.** Patient Characteristics.

| Variables | All participants<br>(N = 282) | Participants<br>without diabetes<br>(N = 150) | Participants<br>with diabetes<br>(N = 132) | P-value |
| --- | --- | --- | --- | --- |
| Age (Mean) | 80.8 ± 6.3 | 81.4 ± 6.1 | 80.1 ± 6.4 | 0.082 |
| Ethnicity |  |  |  |  |
| Malay | 64 (22.7%) | 34 (22.7%) | 30 (22.7%) | 0.338 |
| Chinese | 146 (51.8%) | 84 (56.0%) | 62 (47.0%) |  |
| Indian | 68 (24.1%) | 30 (20.0%) | 38 (28.8%) |  |
| Others | 4 (1.4%) | 2 (1.3%) | 2 (1.5%) |  |
| Female | 162 (57.4%) | 87 (58.0%) | 75 (56.8%) | 0.841 |
| Male | 120 (42.6%) | 63 (42.0%) | 57 (43.2%) |  |
| Smoking | 18 (6.5%) | 11 (7.3%) | 7 (5.4%) | 0.518 |
| Polypharmacy | 115 (40.8%) | 41 (27.3%) | 74 (56.1%) | <0.001 |
| Predischarge CFS<br>score | 6.1 ± 1.1 | 6.0 ± 1.1 | 6.2 ± 1.0 | 0.099 |
| Predischarge CFS<br>score ≥4 | 274 (97.2%) | 144 (96.0%) | 130 (98.5%) | 0.290 |
| Charlson<br>Comorbidity<br>Index | 5.9 ± 1.9 | 5.2 ± 1.7 | 6.7 ± 1.9 | <0.001 |
| History of Stroke | 100 (35.7%) | 48 (32.4%) | 52 (39.4%) | 0.225 |
| Hypertension | 233 (82.6%) | 112 (74.7%) | 121 (91.7%) | <0.001 |
| Dyslipidemia | 121 (42.9%) | 63 (42.0%) | 58 (43.9%) | 0.743 |
| AF | 61 (21.6%) | 31 (20.7%) | 30 (22.7%) | 0.675 |
| CCF | 10 (3.5%) | 3 (2.0%) | 7 (5.3%) | 0.135 |
| IHD | 52 (18.4%) | 17 (11.3%) | 35 (26.5%) | 0.001 |
| PVD | 7 (2.5%) | 3 (2.0%) | 4 (3.0%) | 0.579 |
| CKD (GFR<br><60mL/min) | 107 (37.9%) | 55 (36.7%) | 52 (39.4%) | 0.638 |
| Disability at<br>discharge (MRS<br>≥ 3) | 261 (92.6%) | 135 (90.0%) | 126 (95.5%) | 0.082 |
| Length of stay<br>(days) | 17.6 ± 20.5 | 14.9 ± 11.4 | 20.7 ± 27.1 | 0.018 |
| Discharge<br>Nursing Home | 33 (11.7%) | 14 (9.3%) | 19 (14.4%) | 0.187 |

Data are shown as n (%) for categorical variables, and mean with standard deviation for continuous variables. AF: Atrial Fibrillation, CCF: Congestive Cardiac Failure, IHD: Ischemic Heart Disease, PVD: Peripheral Vascular Disease, CKD: Chronic Kidney Disease, MRS: Modified Rankin Scale

**Table 2.** Prescription rate of cardiovascular medicines in participants with and without diabetes.

| **Medications** | **All participants**<br>**(N=282)** | **Participants without diabetes**<br>**(N= 150)** | **Participants with diabetes**<br>**(N= 132)** | **p-value** |
| --- | --- | --- | --- | --- |
| Statin | 194 (69.0%) | 101 (67.3%) | 93 (71.0%) | 0.508 |
| Antiplatelets | 184 (65.5%) | 93 (62.0%) | 91 (69.5%) | 0.189 |
| CCB (non-DHP & DHP) | 108 (38.4%) | 59 (39.3%) | 49 (37.4%) | 0.740 |
| ACEI/ARB | 55 (19.6%) | 19 (12.7%) | 36 (27.5%) | 0.002 |
| Beta blocker | 53 (18.9%) | 21 (14.0%) | 32 (24.4%) | 0.026 |
| Oral anticoagulants | 34 (12.1%) | 19 (12.7%) | 15 (11.4%) | 0.737 |
| Alpha blocker | 7 (2.5%) | 5 (3.3%) | 2 (1.5%) | 0.332 |
| Spironolactone | 4 (1.4%) | 0 (0%) | 4 (3.1%) | 0.031 |
| Thiazide | 2 (0.7%) | 2 (1.3%) | 0 (0%) | 0.185 |

ACEI: Angiotensin Converting Enzyme Inhibitor, ARB: Angiotensin Receptor Blocker, CCB: calcium channel blocker, DHP: dihydropyridines, Antiplatelet: aspirin, clopidogrel, Oral anticoagulants: warfarin, dabigatran, apixaban, rivaroxaban

In terms of comorbidities, the mean Charlson Comorbidity Index was 5.9 (SD 1.9) in all participants, 5.2 (SD 1.7) in participants without diabetes, and 6.7 (SD 1.9) in participants with diabetes (p<0.001). Participants with diabetes had higher prevalence of hypertension (91.7% vs 74.7% in those without diabetes, p<0.001) and ischemic heart disease (26.5% vs 11.3% in those without diabetes, p=0.001).

Polypharmacy was more common in participants with diabetes (56.1% vs 27.3% in those without diabetes, p <0.001). Participants with diabetes had a significant longer hospitalization, with a mean length of stay of 20.7 days (SD 27.1) vs 14.9 days (SD 11.4) in those without diabetes (p=0.018).

Figure 1 presents the prescriptions of statins and other cardiovascular medicines in the participants. Overall, statin was the most commonly prescribed medication at discharge (69.0%), followed by antiplatelets (65.5%), calcium channel blockers (38.4%), angiotensin converting enzyme inhibitors/angiotensin receptor blockers (19.6%) and beta-blockers (18.9%). There was no significant difference on statin prescription rates between participants with and without diabetes (71.0% vs. 67.3%, respectively, p=0.508).

**Figure 1.**
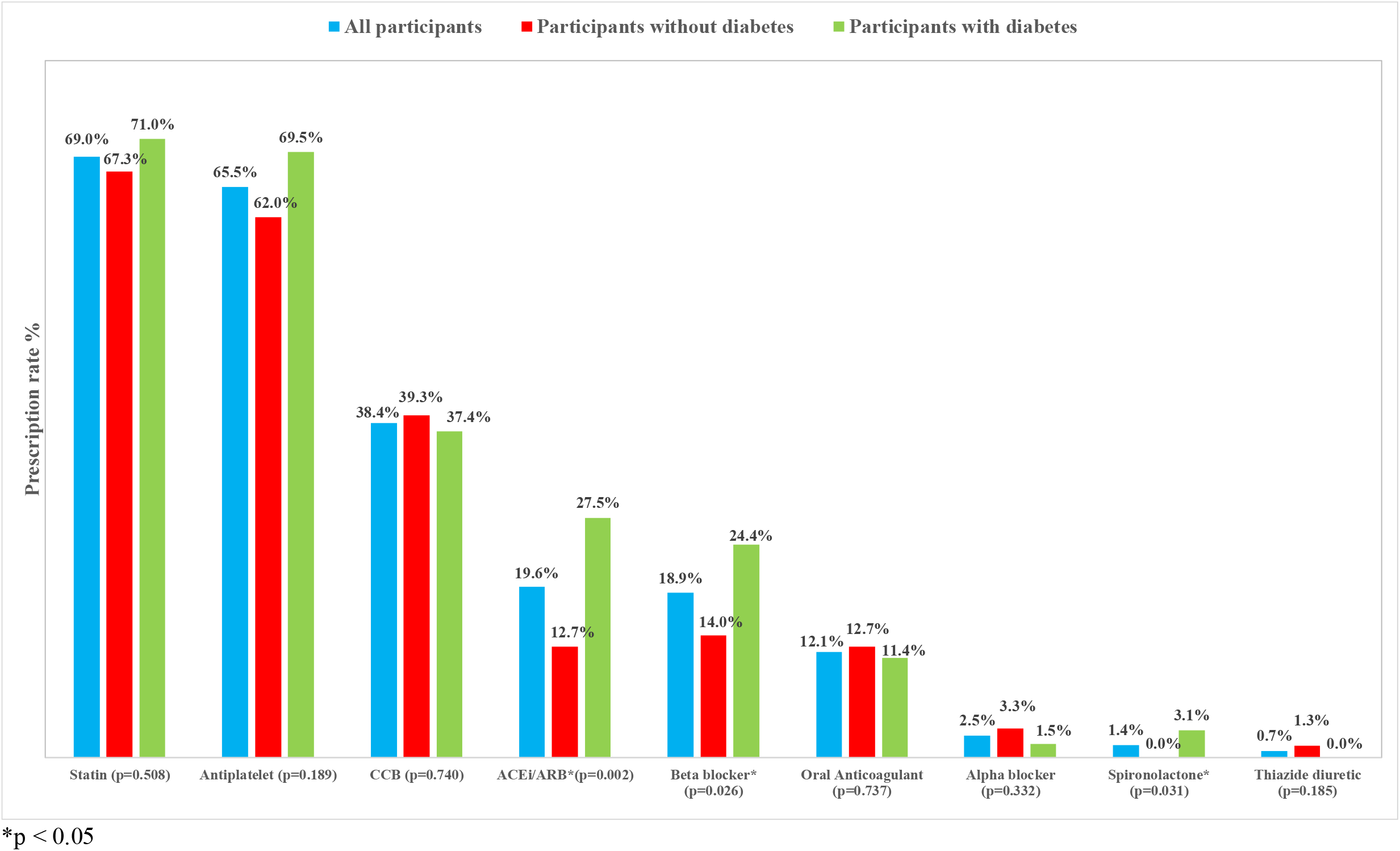
Prescription rates of cardiovascular medications among the participants.

The unadjusted and adjusted odds ratios of frailty and other covariates on statin prescriptions in all participants, without diabetes and with diabetes are presented in Table 3.

**Table 3.** Unadjusted and adjusted odds ratios of frailty and other covariates on statin prescriptions in all participants, without diabetes and with diabetes.

| Variables | All participants |  |  |  | Participants without diabetes |  |  |  | Participants with diabetes |  |  |  |
| --- | --- | --- | --- | --- | --- | --- | --- | --- | --- | --- | --- | --- |
|  | Unadjusted ORs (95%CI) | p | Adjusted ORs (95%CI) | p | Unadjusted ORs (95%CI) | p | Adjusted ORs (95%CI) | p | Unadjusted ORs (95%CI) | p | Adjusted ORs (95%CI) | p |
| CFS score | 0.78<br>(0.61 – 1.02) | 0.065 | 0.64<br>(0.46 – 0.88) | **0.006** | 0.68<br>(0.47 – 0.96) | 0.030 | 0.54<br>(0.33 – 0.89) | **0.016** | 0.94<br>(0.63 – 1.40) | 0.746 | 0.73<br>(0.46 -1.17) | 0.193 |
| Age | 0.97<br>(0.93 – 1.01) | 0.164 | 0.97<br>(0.93 – 1.02) | 0.242 | 0.95<br>(0.89 - 1.00) | 0.057 | 0.93<br>(0.87 – 0.99) | **0.046** | 1.00<br>(0.95 – 1.06) | 0.927 | 1.02<br>(0.95 – 1.10) | 0.530 |
| Male (vs. female) | 0.81<br>(0.49 – 1.34) | 0.410 | 0.68<br>(0.39 – 1.17) | 0.166 | 0.84<br>(0.42-1.67) | 0.617 | 0.51<br>(0.23 – 1.12) | 0.092 | 0.77<br>(0.36 – 1.64) | 0.495 | 0.82<br>(0.35 -1.90) | 0.639 |
| Charlson comorbidity index | 1.03<br>(0.90 – 1.17) | 0.707 | 0.98<br>(0.83 – 1.16) | 0.813 | 0.93<br>(0.77 – 1.14) | 0.493 | 0.98<br>(0.76 – 1.26) | 0.877 | 1.10<br>(0.89 – 1.35) | 0.376 | 0.88<br>(0.67 – 1.17) | 0.381 |
| Dyslipidemia | 1.77<br>(1.04 – 2.99) | 0.034 | 1.86<br>(1.04- 3.32) | **0.036** | 1.38<br>(0.69 – 2.79) | 0.364 | 1.76<br>(0.77 -4.03) | 0.181 | 2.40<br>(1.07 – 5.40) | 0.034 | 3.22<br>(1.20 – 8.69) | **0.021** |
| History of stroke | 1.07<br>(0.63 – 1.81) | 0.814 | 0.99<br>(0.55 – 1.79) | 0.980 | 1.13<br>(0.54 – 2.37) | 0.739 | 1.20<br>(0.51 – 2.83) | 0.669 | 0.97<br>(0.45 – 2.10) | 0.935 | 0.96<br>(0.39 – 2.35) | 0.924 |
| Disability at discharge (MRS ≥ 3) | 1.22<br>(0.47 – 3.17) | 0.686 | 3.75<br>(1.06 – 13.22) | **0.040** | 0.48<br>(0.13-1.8) | 0.279 | 2.22<br>(0.37 -13.26) | 0.381 | 10.82<br>(1.17- 100.29) | 0.036 | 65.40<br>(4.40 – 971.73) | **0.002** |
| Polypharmacy | 1.71<br>(1.01 - 2.92) | 0.047 | 1.86<br>(1.03 – 3.34) | **0.039** | 1.24<br>(0.57 – 2.71) | 0.587 | 1.68<br>(0.69 – 4.11) | 0.255 | 2.28<br>(1.06 – 4.91) | 0.036 | 2.90<br>(1.19 – 7.09) | **0.019** |

On the adjusted models, increased CFS score was significantly associated with reduced odds of receiving statins in all participants (adjusted OR 0.64, 95% CI 0.46 -0.88), and in participants without diabetes (adjusted OR 0.54, 95% CI 0.33 – 0.89), but not in participants with diabetes (adjusted OR 0.73, 95%CI 0.46 – 1.17).

Among the covariates, factors that were significantly associated with statin prescriptions in participants with diabetes included dyslipidemia (adjusted OR 3.22, 95%CI 1.20 – 8.69), disability at discharge (adjusted OR 65.40, 95%CI 4.40 – 971.73), and polypharmacy (adjusted OR 2.90, 95%CI 1.19 – 7.09). In participants without diabetes, the only covariate that was significantly associated with statin prescriptions were age (adjusted OR 0.93, 95%CI 0.87 – 0.99 for each year increase in age).

## Discussion

In our study, only 69.0% of the participants were prescribed statins and frailty was observed to reduce the likelihood of receiving statins in older adults with ischemic stroke. The impact of frailty on the reduced prescription of statins was significant in participants without diabetes compared to those with diabetes.

Our findings are in line with previous studies on statins in patients with strokes. Several studies have reported the underuse of statins in patients post strokes. A study in Finland found that statin was not used by more than 25% of ischemic stroke patients within 90 days after hospital discharge, with women and older patients using statins less frequently.^13^ Another study of 220 older adults with atherosclerotic cardiovascular disease in a tertiary hospital in Saudia Arabia^14^ also reported an under-prescription of statins, with only 59.5% on the therapy. Compared to our study, participants in the Saudi Arabia study were slightly younger (mean age 75 ± 7) and investigators did not include patients with diabetes and further explore impact of frailty.

There are several reasons that may explain the reduced prescription of statins in older patients, particularly in those with frailty. Frail older adults have an inherent risk of polypharmacy.^15^ In our study, polypharmacy was present in more than a third of the study population. As polypharmacy may also sometimes influence adherence and increase the risk of drug-drug interactions, prescribers may prioritize certain medicines over others depending on risk-benefit ratios. This could be done as well to help simplify patients’ medication management plan and improve overall treatment adherence. In addition, there has been some controversy on the use of statins in older people. In older individuals, changes in liver and kidney function can alter drug metabolism and clearance, increasing the risk of adverse effects of statins, such as myopathy, myalgias, muscle weakness, injuries, and cognitive dysfunction.^16^ This could increase the risk of falls^17^, especially in frail older adults who are already at high fall risk. There was also some concern about the potential increased risk of haemorrhagic stroke with statin use.^18^ Even so, two meta-analyses published in 2022 and 2024 reported no increase in the risk of haemorrhagic stroke associated with statins.^19,20^ However, the investigators of these meta-analyses did not take frailty into account in their analyses. In a study of 1665 older Australian men in the Concord Health and Ageing in Men Project^21^, Gnjidic and colleagues found a lack of independent association between the use of statins and institutionalisation or mortality in older and frail men. In a study examining new statin use among older veterans in USA, Orkaby and colleagues also found no significant difference in mortality outcomes between frail and non-frail individuals.^22^ These studies suggest that frailty should not be considered as a “contraindication” for statin use in frail adults.

The absence of a significant association between frailty and statin prescriptions among participants with diabetes may result from the small sample size. However, it may also suggest a difference in prescription patterns among stroke patients with and without diabetes. In our study, among participants without diabetes, age and frailty were the only factors that were associated with statin prescriptions: advanced age and higher levels of frailty reduced the odds of receiving statins. In contrast, these two factors did not significantly affect statin prescriptions among participants with diabetes, while factors such as dyslipidemia and the consequences of strokes (as reflected by disability at discharge) may have a stronger influence on clinicians’ decisions of prescribing. Diabetes is a risk factor for cardiovascular events, hence post-stroke older patients with diabetes require careful management compared to those without this condition. There has been evidence that the lack of statins early after ischemic stroke was associated with increased probability of poor health outcomes.^13^ Whether this is true for frail older adults is yet to be seen. Further studies are needed to understand the perspectives of clinicians in prescribing statins for stroke prevention in older individuals with frailty and diabetes post ischemic strokes. This understanding can ultimately guide more tailored and effective management strategies for this vulnerable group.

## Strength and limitation

To the best of our knowledge, our study is the first to show the influence of frailty on statin prescription and the potential interaction between frailty and diabetes in older Malaysians with ischemic stroke. However, this study has some limitations. Data of serum cholesterol levels and medication use prior to admission were not obtained. Additionally, there was no information available regarding factors that could be contraindications for statin use, such as abnormal liver function or a previous history of adverse effects. Due to the cross-sectional nature of study, we were unable to explore the impact of these interactions on adverse outcomes such as re-hospitalization or mortality. The study was conducted at a single tertiary hospital and findings may not be representative of all older patients with stroke in Malaysia.

## Conclusion

This study in older Malaysians post ischemic stroke showed that frailty was associated with reduced odds of receiving statins. The differences in the relationship between frailty and statin prescription among participants with and without diabetes may suggest a personalized approach in secondary prevention for older patients after strokes. Future studies should include longitudinal outcomes to help clarify the usefulness of statin in secondary prevention post ischemic stroke, particularly among frail older adults with diabetes. Prescribers’ perspectives could also be sought to better understand risk-benefit consideration, aiding in the development of personalized healthcare for older individuals.

## Data Availability

All data produced in the present study are available upon reasonable request to the authors

